# Immunotoxic Effects in Children Resulting from Prenatal and Early Childhood Exposure to Pesticides: Protocol and Pilot Study of a Systematic Review and Meta-Analysis

**DOI:** 10.1101/2024.09.06.24313180

**Authors:** Moustafa Sherif, Aya Darwish, Balázs Ádám

**Affiliations:** Institute of Public Health, College of Medicine and Health Sciences, United Arab Emirates University, POBox 15551, Al-Ain, AbuDhabi, United Arab Emirates; Department of Physiology, Faculty of Veterinary Medicine, Benha University, POBox 17321, Toukh, Al-Qalyubia, Egypt

**Keywords:** Pesticides, immunotoxicity, children, prenatal exposure, vulnerable periods

## Abstract

**Background:** Pesticides, commonly employed in both agricultural and domestic environments, have been associated with disturbing the balance of the immune system during prenatal and early childhood phases of development. This disturbance has the potential to result in various outcomes such as autoimmunity, immunosuppression, inflammation, hypersensitivity, tissue damage, and overall alterations in immune modulation. The escalating concern regarding the immunotoxic effects of pesticides necessitates a systematic review and potential meta-analysis to consolidate empirical evidence and address the knowledge gaps. This systematic review aims to comprehensively assess the immunotoxic impact of pesticide exposure on children, considering alterations in immune system function induced by specific types of pesticides.

**Methods:** Through a meticulous search of the literature and synthesis of findings, the review will scrutinize prevalence, effect size, mechanisms, vulnerable periods, and the role of pre-existing conditions. The submitted protocol to PROSPERO aligns with the Preferred Reporting Items for Systematic Reviews and Meta-Analyses Protocols (PRISMA-P) statement, ensuring methodological rigor. A preliminary exploration involved a systematic search in electronic databases based on a pre-defined Population, Exposure, Comparator, and Outcome (PECO) statement. Data extraction sheets and risk of bias assessment tools were piloted on eight eligible articles to ensure reliability and efficiency. Subsequently, a rerun of the search will be executed, and two independent reviewers will employ the systematic review tool Covidence for article screening and data extraction, and Excel for risk of bias assessment. The results will be synthetized narratively in summary tables, and, if findings allow, meta-analysis including subgroup and sensitivity analysis will be conducted.

**Discussion:** By elucidating the intricate relationship between pesticide exposure and immunotoxicity during prenatal and early childhood, this review will provide insights into immune response that can contribute to public health awareness and guide interventions.

**PROSPERO Registration number:** [CRD42024510916].

**Impact Statement:** This study will establish a high level of evidence to highlight the critical immunotoxic risks of prenatal and early childhood pesticide exposure, offering essential insights for public health policies and interventions to safeguard the infants during this vulnerable developmental periods.

## Background

Human susceptibility to environmental factors, particularly pesticides, has emerged as a significant public health concern ^1,2^. The Developmental Origin of Human Disease (DOHAD) theory, as proposed by David Barker, asserts that environmental conditions experienced in the initial stages of development can have lasting effects on subsequent phases of life ^3^. Consequently, exposure to environmental pollutants during the in-utero period can increase the predisposition to a range of pathologies, manifesting in both early and later stages of life. Recognizing and addressing the impact of environmental elements on human health across developmental stages becomes imperative in light of these findings. An increasing body of evidence suggests that children’s health is profoundly influenced by exposure to pesticides that are widely used in both agricultural and household settings. These chemical compounds can have significant consequences when interfering with embryonic development ^4^.

Women, especially who are living in areas with spraying activities to control vector borne diseases, face significant exposure to organochlorine (OC) pesticides like DDT and its byproducts. Studies show high levels of these pesticides in maternal serum (26.2 µg/g lipids) and cord blood (up to 4.4 µg/g lipids), indicating their ability to pass through the placenta. Breast milk further exposes infants, with concentrations reaching 6.9 µg/g lipids ^5^. Susceptibility during prenatal and early childhood is attributed to global epigenetic reprogramming, increased rates of cell proliferation, and intricate organogenesis events ^6^. Various factors, including the quantity, timing, and nature of exposure, play a significant role in shaping long-term immune responses, that may also be influenced by genetic factors, as previously indicated ^7,8^. Aligning with the exposome paradigm ^9^, it is crucial to take into account the cumulative impact of a wide range of environmental factors when assessing the effects of prenatal exposure on the immune system.

Few studies investigated the impact of pesticides and other environmental contaminants on infants’ immune responses, revealing different levels and directions of associations. For instance, a study examined the impact of exposure to dioxins, PCBs, and organochlorine pesticides on 10-month-old infants revealed that higher concentrations of heptachlor epoxide, chlordane, and dioxins in breast milk were significantly associated with increased percentages of CD8+ T cells and CD3+ T cells, along with alterations in the CD4+-to-CD8+ ratio ^10^. Furthermore, T cells isolated from cord blood exhibited a negative association between their proliferation, when stimulated, and levels of PCBs and 4,41-dichlorodiphenyl-dichloroethene (4,41-DDE) in newborns prenatally exposed to organochlorine compounds and mercury ^11^. Another finding indicated that the secretion of TNF-α by cord blood mononuclear cells, under mitogen stimulation, was negatively associated with prenatal organochlorine exposure ^12^. In 6-month-old infants, elevated levels of PCB-153 and DDE were significantly linked to lower levels of BCG-specific antibodies ^13^. Notably, a study ^14^ discovered higher pesticide levels in umbilical cord plasma than in mothers, but genotoxicity markers in immune cells did not correlate. This lack of correlation might be attributed to variations in exposure, pregnancy-related processes, and individual differences.

The mechanisms of effects of prenatal exposures on the foetal immune system have numerous knowledge gaps because various pesticides can have different impacts based on the organ, cell type, and molecular pathway ^15–17^, which is necessitating the consolidation of the current evidence to highlight further considerations of research for the future. For example, there are no specific validated biomarkers in tissues, such as in placenta or body fluids, that can be used to capture exposure-dependent differences in changes in the immune system ^18^.

## Objective

By synthesizing existing literature, this systematic review with a potential meta-analysis aims to critically evaluate the immunotoxic consequences arising from pesticide exposure during crucial developmental phases, showcasing a nuanced interplay that can shift immune equilibrium towards immunosuppression or immunoenhancement, heightening the risk for infectious or chronic diseases, hypersensitivity, allergies, inflammation, tissue injury, or autoimmunity. It will consolidate empirical evidence, provide valuable insights and address knowledge gaps in the intricate relationship between pesticide exposure and immunotoxicity in children. Understanding the complex pathways and mechanisms influenced by prenatal immunotoxic exposures necessitates a systematic approach, with a thorough evaluation of the specific pesticides and their metabolites. This systematic review will provide synthesized knowledge that can guide researchers to develop targeted intervention strategies, by highlighting the numerous epidemiological studies linking pesticide exposure to alterations in children’s immune responses. Recognizing the sources and routes of exposure during critical periods is crucial to unravel the complex pathways through which pesticides exert immunotoxic effects. Additionally, identifying commonly used biomarkers and endpoints for assessing immune system effects enhances methodological robustness.

## Methods/Design

### 1. Protocol and registration

The protocol was registered in the International Prospective Register of Systematic Reviews (PROSPERO) [CRD42024510916]. This protocol follows the PRISMA-P statement that refers to the Preferred Reporting Items for Systematic Reviews and Meta-Analyses Protocols ^19^. The scheduled start date of the study is 1st April 2023 and will continue until 1st November 2024. The PRISMA-P checklist is accessible as a supplemental file (S1). The final review will go along with the new PRISMA 2020 statement ^20,21^.

### 2. Eligibility criteria

A population, exposure, comparator and outcomes (PECO) statement was established to review data on immunotoxicity of pesticide exposures occurring in the prenatal and postnatal period to infants and children below five years of age.

#### A. Participants/population

Inclusion: Embryos, fetuses, infants, young children up to 5 years of age, and pregnant mothers (if the placenta was investigated) will be considered.

Exclusion: Children over 5 years of age and pregnant mothers who developed immunotoxicity in any tissue other than the placenta will be excluded.

#### B. Intervention(s), exposure(s)

Inclusion: Exposure to pesticides during the prenatal and/or early childhood periods up to the age of 5 years, regardless of whether they provide specific information on the types of pesticides or their metabolites or the route of exposure, as well as biomarkers of pesticide exposure in mothers to draw associations with the level of immunotoxicity in their paired children, will be considered. Additionally, exposure through maternal blood, placental transfer, or breastfeeding will be taken into account.

Exclusion: Exposure to environmental contaminants other than pesticides and exposure to pesticides during time periods other than the prenatal and/or early childhood periods.

#### C. Comparator/Control

A comparison group (e.g., unexposed or low-exposure group) are required specifically for assessing the association/effect size. No comparator group is needed for research on prevalence of exposure, prevalence and mechanisms of effects. The mothers can act as comparator for their paired children.

#### D. Outcome

The systematic review will include studies reporting outcomes specifically related to the immunotoxic impact of prenatal and early childhood pesticide exposure in children. Eligible outcomes for inclusion encompass immune system function, incorporating measures such as cytokine profiles, immune cell counts, and immune cell activity, as well as associations between prenatal/early childhood pesticide exposure and immunotoxicity. Studies reporting outcomes outside of these predefined categories or specifically focusing on allergies, infectious diseases, and childhood cancers, including leukemia and lymphoma, will be excluded from the review. This exclusion aims to maintain a focused investigation specifically on the impact of pesticide exposure on the functioning of the immune system, without extending immunotoxicity to particular health conditions.

### 3. Types of studies

The review will encompass observational studies, including cross-sectional, case-control, and cohort studies, to investigate the research objectives. Conversely, certain types of studies will be excluded from the review, such as *in vitro* and *in vivo* animal studies, case reports, opinion articles, commentaries, letters, review articles, published abstracts, and conference proceedings. By adhering to these inclusion and exclusion criteria, the review aims to focus on high-quality original studies that directly contribute to addressing the research objectives.

### 4. Information sources

We will search systematically for studies that match our criteria and are published in peer-reviewed scientific journals in electronic four databases: PubMed (NLM), EMBASE (Elsevier), Web of Science (Clarivate), and Scopus (Elsevier).

### 5. Search strategy

We conducted preliminary searches in the four databases in February 2024 based on the PECO statement with inclusion and exclusion criteria as defined above. Identified search terms were systematically developed with the help of PubMed’s MeSH and the search technical specifications. The pre-search is available in a supplemental file (S2).

A full systematic search of the literature will be conducted in April 2024 based on the strategy developed in the pre-search. The search will be performed by applying all search terms in the databases MeSH/thesauruses and in the “title” and “abstract” fields. All databases will be searched from their inception. A filter for English language will be used. No additional filers or limitations will be applied for the best possible results and to capture all pre-indexed studies. The PRISMA S checklist for searching and reproducible search strings for all databases will be provided as an appendix in the final review.

The search will be updated in all databases ahead of completing the study to ensure the inclusion of potential eligible studies published during the review process. Finally, the reference lists of the included studies and identified systematic reviews will be screened manually by two independent reviewers.

### 6. Study records and management

#### A. Data management

Records located in the literature search will be uploaded to the systematic review software Covidence for title/abstract and full text screening, conflict resolving, selection, and data extraction. All modules in Covidence are blinded.

#### B. Selection process

All records retrieved in the search will be automatically de-deduplicated and screened in Covidence. Two review authors will independently screen the titles and abstracts of the studies and assess them if they are potentially eligible according to the inclusion and exclusion criteria. The same reviewers together with the National Medical Library team at UAEU will then work to recover the full text of the selected studies. If there are any disagreements about inclusion of certain studies in either screening rounds, consensus will be determined by a third reviewer through the blinded conflict module in Covidence. PRISMA flow-diagram will be used to record the results of the screening and selection process, including reasons for full text exclusion ^21^. Finally, Cabell’s Predatory Reports will be used to confirm the academic quality of eligible studies published in open access journals.

### 7. Data collection process

The data extraction template was developed and pilot tested on eight articles in Excel as shown in the supplemental file (S3). The template will be populated in Covidence, where independent data extraction will be performed by two reviewers. In case of any discrepancies, a third reviewer will resolve it using the blinded conflict resolving function of the extraction module in Covidence. If necessary, we will contact study authors to retrieve any missing data.

### 8. Data items and extraction

Data extraction will include publication details, specifics of study settings, in-depth characteristics of the study populations and their sample sizes, demographic information regarding the participants and their baseline characteristics, descriptions of the types of pesticides to which participants are exposed, details about the levels of pesticide exposure (measured or assessed), biomarkers and endpoints of immunotoxicity, effect sizes and strength of associations. Additionally, information will be retrieved on the identified mechanisms by which pesticide exposure during the prenatal and early childhood periods may affect immune system development in children. Finally, information on conflicts of interest and funding sources will also be collected. If uncertainties arise, the data extractor will initiate a discussion with a second reviewer to make the final decision.

### 9. Risk of bias (RoB) in individual studies

The risk of bias in the included studies will be appraised by two skilled reviewers using the Newcastle-Ottawa Scale (NOS) for case-control and cohort studies. Additionally, based on NOS, we have developed a novel risk of bias tool for assessing bias in cross-sectional studies. In cases where there are uncertainties regarding the risk of bias for specific studies, a third reviewer will be consulted to resolve it. The RoB tools, including the newly developed for this systematic review, were pilot tested on eight articles as displayed in supplemental file (S4). Two reviewers will individually assess the RoB in the eligible studies. Any discrepancies over the risk of bias assessment of particular studies between the two reviewers will be solved by a third reviewer.

### 11. Data synthesis

We will perform a comprehensive narrative synthesis of the findings extracted from the included studies, structured around the type of pesticides under investigation, population characteristics, and the nature of immunotoxicity. This synthesis will provide a coherent and systematic overview of the results and conclusions derived from the primary research.

If two or more studies are found with appropriate estimates on outcome frequency and/or on effect size, two reviewers will individually explore the clinical heterogeneity of the studies and combine them for meta-analysis. A meta-analysis may be conducted specifically to quantify the effect size of prenatal/early childhood pesticide exposure immunotoxicity in children. In addition to the narrative synthesis, this quantitative analysis will provide a more precise assessment of the strength and direction of the association between pesticide exposure and immunotoxicity. The decision to conduct a meta-analysis will be based on the availability and quality of data from the selected studies, and it will be detailed in the systematic review.

If the combined studies are clinically sufficiently homogenous, the frequency and/or effect estimates will be pooled in a quantitative meta-analysis, using the inverse variance method with a random-effects model to account for cross-study heterogeneity ^22^. Statistical heterogeneity of the studies will be analyzed using the I2 statistics ^23^. The meta-analysis will be performed in RevMan software ^24^. The combined estimations will be shown in forest plots.

The funnel plot graphic representation will be used to visually evaluate publication bias, and we will run a sensitivity analysis in case of the presence of outliers or asymmetry in the funnel plot.

### 12. Analysis of subgroups or subsets

Analysis of subgroups or subsets will be conducted if there are indications of variations in the strength of association and effect size of immune system impairment estimates based on population characteristics, such as country, sex, age, on type of pesticides or their specific metabolites, as well as on the mechanisms by which they may impact the immune system. These subgroup analyses will allow for a more detailed examination of potential differences within specific subgroups, providing valuable insights into the relationship between pesticide exposure and its impact on immunotoxicity in children.

### 13. Quality of cumulative evidence (QoE)

The quality of evidence will be assessed by all reviewers using the Grading of Recommendations Assessment, Development and Evaluation (GRADE) tool to assess individual risk for bias, inconsistency, indirectness, imprecision, and publication bias. GRADE scoring assessment tool will allow us to provide summary-based evidence statements ^25,26^.

## Discussion

### 1. Purpose

A successful pregnancy relies on effective communication between the mother and the developing baby. The placenta is vital for providing nutrients, supporting hormone functions, and processing medications during the embryo’s growth ^27^. This communication includes not only normal substances but also potential dangers like drugs, food additives, pesticides, and environmental pollutants ^28^. Under normal conditions, a healthy immune system physiologically maintains a balance between its innate and adaptive components, distinguishing self from nonself and combating foreign material; however, environmental or occupational exposures can disrupt this balance, leading to immune-suppression or -enhancement and therefore increase in vulnerability to infectious or chronic diseases, respectively ^29^. This systematic review aims to comprehensively analyze the immunotoxic impact of prenatal and early childhood exposure to pesticides in children, with defined objectives to evaluate pesticide impact and identify vulnerable developmental periods through synthesizing existing literature and addressing knowledge gaps. It will assess the prevalence of pesticide exposure during critical developmental periods, determine the association with immunotoxicity, and identify specific pesticides and metabolites impacting the immune system. The study will investigate mechanisms of immunotoxicity, identifying biomarkers and endpoints in prenatal stages and in children up to five years of age. Additionally, it will examine available evidence on pre-existing conditions and vulnerable periods influencing susceptibility. Through meticulous application of inclusion and exclusion criteria and systematic search strategies, the study provides insights for researchers and policymakers, contributing to the understanding of immunotoxic consequences and informing public health initiatives.

### 2. Strengths and limitations

Limitations of this study include potential challenges in uncovering pathomechanisms due to the observational nature of the included studies. While analytical epidemiological studies can establish causality, their capacity to delve into detailed pathomechanisms is typically limited. The diversity in study designs, exposure assessment methods, and outcome measurements across different studies may introduce heterogeneity, limiting the ability to conduct a uniform meta-analysis. Additionally, the reliance on published literature may imply publication bias, as negative or non-significant findings may be underrepresented. The lack of standardized biomarkers for assessing immune system effects and the variability in the timing and duration of pesticide exposure in the included studies may also pose challenges in synthesizing consistent evidence. Moreover, the available evidence may be influenced by confounding factors, such as genetic predispositions and other environmental exposures, that were not consistently accounted for across studies.

Despite these limitations, the study has several strengths. The systematic review adopts a rigorous and transparent methodology, aligning with the PRISMA-P guidelines and utilizing established risk of bias assessment tools. The plans for a potential meta-analysis enhance the quantitative synthesis of evidence, providing a more robust evaluation of the association between pesticide exposure and immunotoxicity. The comprehensive search strategy, involving major electronic databases and manual screening of reference lists, ensures a thorough exploration of the existing literature. The focus on vulnerable populations, such as pregnant mothers and children up to five years of age, contributes to the identification of critical developmental periods.

### 3. Implications for practice and research

By addressing knowledge gaps and consolidating empirical evidence, this study offers valuable insights for researchers, policymakers, and public health initiatives seeking to understand and mitigate the immunotoxic consequences of pesticide exposure early in life.

## Data Availability

All data produced in the present work are contained in the manuscript

https://alumniuaeuac-my.sharepoint.com/:f:/g/personal/202190149_uaeu_ac_ae/Ev4tse1Kc7pAl6ui4V7lrcsBzCbpnIT8dyW3MTeo7CXieA?e=IRRQYe

## List of abbreviations

PRISMA: Preferred Reporting Items for Systematic Reviews and Meta-analyses
PECO: population, exposure, comparator and outcomes
RoB: Risk of bias
NOS: Newcastle-Ottawa Scale
GRADE: Grading of Recommendations Assessment, Development and Evaluation
QoE: Quality of cumulative evidence

## Supplementary Information

The online version contains supplementary material available at: XXX

PRISMA-P checklist: supplemental file (S1)

Pre-search string: supplemental file (S2)

Extraction sheet including pilot extraction: supplemental file (S3)

RoB sheet including pilot RoB assessment: supplemental file (S4)

## Acknowledgements

Not applicable

## Authors’ contributions

MS, AD, and BÁ were involved in conceptualization and protocol development. MS and AD have developed the search strategy. Literature search will be conducted by MS and AD. Screening will be performed by MS and AD, with BÁ resolving conflicts. MS and AD will extract data, which will be validated by BÁ. RoB will be performed by MS and AD, with BÁ resolving conflicts. Quality of evidence will be assessed by all reviewers.

## Availability of data and materials

Data sharing is not applicable to this article as no datasets were generated or analysed during the current study.

## Declarations

### Ethics approval and consent to participate

Ethics approval is not necessary for such type of research (systematic review), because no public members or patients will participate in the study. No patient consent is required for this study.

### Consent for publication

Not applicable

## References

1. Ramadhan Makame K, Sherif M, Ostlundh L, Sandor J, Adam B, Nagy K. Are encapsulated pesticides less harmful to human health than their conventional alternatives? Protocol for a systematic review of in vitro and in vivo animal model studies. Environ Int. 2023;174:107924. doi:10.1016/j.envint.2023.107924

2. Sherif M, Makame KR, Ostlundh L, et al. Genotoxicity of Occupational Pesticide Exposures among Agricultural Workers in Arab Countries: A Systematic Review and Meta-Analysis. Toxics. 2023;11(8):663. doi:10.3390/toxics11080663

3. Wadhwa PD, Buss C, Entringer S, Swanson JM. Developmental origins of health and disease: brief history of the approach and current focus on epigenetic mechanisms. Semin Reprod Med. 2009;27(5):358–368. doi:10.1055/s-0029-1237424

4. Tang ZR, Xu XL, Deng SL, Lian ZX, Yu K. Oestrogenic Endocrine Disruptors in the Placenta and the Fetus. Int J Mol Sci. 2020;21(4):E1519. doi:10.3390/ijms21041519

5. al-Saleh I, Shinwari N, Basile P, et al. DDT and its metabolites in breast milk from two regions in Saudi Arabia. J Occup Environ Med. 2003;45(4):410–427. doi:10.1097/01.jom.0000058344.05741.22

6. Legoff L, D’Cruz SC, Tevosian S, Primig M, Smagulova F. Transgenerational Inheritance of Environmentally Induced Epigenetic Alterations during Mammalian Development. Cells. 2019;8(12):E1559. doi:10.3390/cells8121559

7. Chen CH, Lee YL, Wu MH, et al. Environmental tobacco smoke and male sex modify the influence of IL-13 genetic variants on cord blood IgE levels. Pediatr Allergy Immunol Off Publ Eur Soc Pediatr Allergy Immunol. 2012;23(5):456–463. doi:10.1111/j.1399-3038.2012.01278.x

8. Fedulov AV, Kobzik L. Allergy risk is mediated by dendritic cells with congenital epigenetic changes. Am J Respir Cell Mol Biol. 2011;44(3):285–292. doi:10.1165/rcmb.2009-0400OC

9. Hong X, Liu C, Chen X, et al. Maternal exposure to airborne particulate matter causes postnatal immunological dysfunction in mice offspring. Toxicology. 2013;306:59–67. doi:10.1016/j.tox.2013.02.004

10. Nagayama J, Tsuji H, Iida T, et al. Immunologic effects of perinatal exposure to dioxins, PCBs and organochlorine pesticides in Japanese infants. Chemosphere. 2007;67(9):S393–398. doi:10.1016/j.chemosphere.2006.05.134

11. Belles-Isles M, Ayotte P, Dewailly E, Weber JP, Roy R. Cord blood lymphocyte functions in newborns from a remote maritime population exposed to organochlorines and methylmercury. J Toxicol Environ Health A. 2002;65(2):165–182. doi:10.1080/152873902753396794

12. Bilrha H, Roy R, Moreau B, Belles-Isles M, Dewailly E, Ayotte P. In vitro activation of cord blood mononuclear cells and cytokine production in a remote coastal population exposed to organochlorines and methyl mercury. Environ Health Perspect. 2003;111(16):1952–1957. doi:10.1289/ehp.6433

13. Jusko TA, De Roos AJ, Lee SY, et al. A Birth Cohort Study of Maternal and Infant Serum PCB-153 and DDE Concentrations and Responses to Infant Tuberculosis Vaccination. Environ Health Perspect. 2016;124(6):813–821. doi:10.1289/ehp.1510101

14. Alvarado-Hernandez DL, Montero-Montoya R, Serrano-Garcia L, Arellano-Aguilar O, Jasso-Pineda Y, Yanez-Estrada L. Assessment of exposure to organochlorine pesticides and levels of DNA damage in mother-infant pairs of an agrarian community. Environ Mol Mutagen. 2013;54(2):99–111. doi:10.1002/em.21753

15. Farzan SF, Korrick S, Li Z, et al. In utero arsenic exposure and infant infection in a United States cohort: a prospective study. Environ Res. 2013;126:24–30. doi:10.1016/j.envres.2013.05.001

16. Rahman A, Vahter M, Ekstrom EC, Persson LA. Arsenic exposure in pregnancy increases the risk of lower respiratory tract infection and diarrhea during infancy in Bangladesh. Environ Health Perspect. 2011;119(5):719–724. doi:10.1289/ehp.1002265

17. Raqib R, Ahmed S, Sultana R, et al. Effects of in utero arsenic exposure on child immunity and morbidity in rural Bangladesh. Toxicol Lett. 2009;185(3):197–202. doi:10.1016/j.toxlet.2009.01.001

18. Rychlik KA, Sille FCM. Environmental exposures during pregnancy: Mechanistic effects on immunity. Birth Defects Res. 2019;111(4):178–196. doi:10.1002/bdr2.1469

19. Moher D, Shamseer L, Clarke M, et al. Preferred reporting items for systematic review and meta-analysis protocols (PRISMA-P) 2015 statement. Syst Rev. 2015;4:1. doi:10.1186/2046-4053-4-1

20. Cumpston M, Li T, Page MJ, et al. Updated guidance for trusted systematic reviews: a new edition of the Cochrane Handbook for Systematic Reviews of Interventions. Cochrane Database Syst Rev. 2019;10:ED000142. doi:10.1002/14651858.ED000142

21. Page MJ, McKenzie JE, Bossuyt PM, et al. The PRISMA 2020 statement: an updated guideline for reporting systematic reviews. BMJ. 2021;372:n71. doi:10.1136/bmj.n71

22. Woodruff TJ, Sutton P. The Navigation Guide systematic review methodology: a rigorous and transparent method for translating environmental health science into better health outcomes. Environ Health Perspect. 2014;122(10):1007–1014. doi:10.1289/ehp.1307175

23. Higgins JPT, Thompson SG, Deeks JJ, Altman DG. Measuring inconsistency in meta-analyses. BMJ. 2003;327(7414):557–560. doi:10.1136/bmj.327.7414.557

24. Schmidt L, Shokraneh F, Steinhausen K, Adams CE. Introducing RAPTOR: RevMan Parsing Tool for Reviewers. Syst Rev. 2019;8(1):151. doi:10.1186/s13643-019-1070-0

25. Balshem H, Helfand M, Schunemann HJ, et al. GRADE guidelines: 3. Rating the quality of evidence. J Clin Epidemiol. 2011;64(4):401–406. doi:10.1016/j.jclinepi.2010.07.015

26. Morgan RL, Thayer KA, Bero L, et al. GRADE: Assessing the quality of evidence in environmental and occupational health. Environ Int. 2016;92-93:611–616. doi:10.1016/j.envint.2016.01.004

27. Sastry. Techniques to study human placental transport. Adv Drug Deliv Rev. 1999;38(1):17–39. doi:10.1016/s0169-409x(99)00004-628.

28. Myllynen P, Pasanen M, Pelkonen O. Human placenta: a human organ for developmental toxicology research and biomonitoring. Placenta. 2005;26(5):361–371. doi:10.1016/j.placenta.2004.09.006

29. Lehmann I. [Environmental pollutants as adjuvant factors of immune system derived diseases]. Bundesgesundheitsblatt Gesundheitsforschung Gesundheitsschutz. 2017;60(6):592–596. doi:10.1007/s00103-017-2545-6

