## Supplemental file S2 for "Immunotoxic Effects in Children Resulting from Prenatal and Early Childhood Exposure to Pesticides: Protocol and Pilot Study of a Systematic Review and Meta-Analysis"

| **Database** | **Search Terms** | **Hits and date** |
| --- | --- | --- |
| Pubmed | (("Immun*"[Title/Abstract] OR "Antibody-Producing Cell*"[Title/Abstract] OR "Lymphocyte*"[Title/Abstract] OR "Antigen-Presenting Cell*"[Title/Abstract] OR "Dendritic Cell*"[Title/Abstract] OR "Dendritic Cell*"[Title/Abstract] OR "Bone Marrow*"[Title/Abstract] OR "Enterochromaffin*"[Title/Abstract] OR "Immunological Synapse*"[Title/Abstract] OR "Leukocyte*"[Title/Abstract] OR "Granulocyte*"[Title/Abstract] OR "Basophil*"[Title/Abstract] OR "Eosinophil*"[Title/Abstract] OR "Leukocyte*"[Title/Abstract] OR "Mononuclear*"[Title/Abstract] OR "Lymph*"[Title/Abstract] OR "Mast Cell*"[Title/Abstract] OR "T-cell*"[Title/Abstract] OR "T cell*"[Title/Abstract] OR "Phagocyte*"[Title/Abstract] OR "Macrophage*"[Title/Abstract] OR "Monocyte*"[Title/Abstract] OR "cytokine*"[Title/Abstract] OR "Neutrophil*"[Title/Abstract] OR "B-cell*"[Title/Abstract] OR "B cell*"[Title/Abstract] OR Lymphangiogene*[Title/Abstract] OR "antibod*"[Title/Abstract] OR "autoimmun*"[Title/Abstract] OR "major histocompat*"[Title/Abstract] OR "Hapteni*"[Title/Abstract] OR "Immune System"[Mesh] OR "Immune System Diseases"[Mesh]) AND ("agrochemical*"[Title/Abstract] OR "agrichemical*"[Title/Abstract] OR "plant protection product*"[Title/Abstract] OR "pesticid*"[Title/Abstract] OR "biocid*"[Title/Abstract] OR "herbicid*"[Title/Abstract] OR "weedkiller*"[Title/Abstract] OR "weed killer*"[Title/Abstract] OR "defolian*"[Title/Abstract] OR "insecticid*"[Title/Abstract] OR "nematicid*"[Title/Abstract] OR "molluscicid*"[Title/Abstract] OR "piscicid*"[Title/Abstract] OR "avicid*"[Title/Abstract] OR "rodenticid*"[Title/Abstract] OR "repellent*"[Title/Abstract] OR "antiparasit*"[Title/Abstract] OR "lampricid*"[Title/Abstract] OR "acaricid*"[Title/Abstract] OR "miticid*"[Title/Abstract] OR "mite control*"[Title/Abstract] OR "algicid*"[Title/Abstract] OR "algaecid*"[Title/Abstract] OR "chemosterilant*"[Title/Abstract] OR "Agrochemicals"[MeSH:NoExp] OR "Pesticides"[MeSH])) AND ("Infant*"[Title/Abstract] OR "Prenatal*"[Title/Abstract] OR "postnatal*"[Title/Abstract] OR "antenatal*"[Title/Abstract] OR "Embryonic and Fetal Development"[Mesh] OR "Embryo*" [Title/Abstract] OR "Fetal*"[Title/Abstract] OR "foetal*"[Title/Abstract] OR "foetus*"[Title/Abstract] OR "fetus*"[Title/Abstract] OR "Gestational Age*"[Title/Abstract] OR "Infant"[Mesh] OR "Prenatal Diagnosis"[Mesh]) | 1,298 in 09/02/2024 |
| Web of science | **"**Infant*" OR "Prenatal*" OR “postnatal*” OR “antenatal*” OR "embryo*” OR “fetal*” OR “foetal*” OR “foetus*” OR “fetus*” OR “Gestation*” (Topic) and “agrochemical*” OR “agrichemical*” OR "plant protection product*" OR “pesticid*” OR “biocid*” OR “herbicid*” OR “weedkiller*” OR "weed killer*" OR “defolian*” OR “insecticid*” OR “nematicid*” OR “molluscicid*” OR “piscicid*” OR “avicid*” OR “rodenticid*” OR “repellent*” OR "antiparasit*" OR “lampricid*” OR “acaricid*” OR “miticid*” OR "mite control*" OR “algicid*” OR “algaecid*” OR “chemosterilant*” (Topic) and “Immun*” OR “Antibody-Producing Cell*” OR “Lymphocyte*” OR “Antigen-Presenting Cell*” OR “Dendritic Cell*” OR “Bone Marrow*” OR “Enterochromaffin*” OR “Immunological Synapse*” OR “Leukocyte*” OR “Granulocyte*” OR “Basophil*” OR “Eosinophil*” OR “Leukocyte*” OR “Mononuclear*” OR “Lymph*” OR “Mast Cell*” OR “Phagocyte*” OR “Macrophage*” OR “Monocyte*” OR “Neutrophil*” OR “B cell*” OR “B-cell*” OR “Lymphangiogene*” OR “T cell*” OR “T-cell*” OR “Cytokine*” OR “innate*” OR “autoimmun*” OR “antibod*” OR “Major histocomp*” OR “Hapteni*” (Topic) | 1,086 in 09/02/2024 |
| Scopus | **( TITLE-ABS-KEY ( "Infant*" OR "Prenatal*" OR "postnatal*" OR "antenatal*" OR "embryo*" OR "fetal*" OR "foetal*" OR "foetus*" OR "fetus*" OR "Gestation*" ) AND TITLE-ABS-KEY ( "agrochemical*" OR "agrichemical*" OR "plant protection product*" OR "pesticid*" OR "biocid*" OR "herbicid*" OR "weedkiller*" OR "weed killer*" OR "defolian*" OR "insecticid*" OR "nematicid*" OR "molluscicid*" OR "piscicid*" OR "avicid*" OR "rodenticid*" OR "repellent*" OR "antiparasit*" OR "lampricid*" OR "acaricid*" OR "miticid*" OR "mite control*" OR "algicid*" OR "algaecid*" OR "chemosterilant*" ) AND TITLE-ABS-KEY ( "Immun*" OR "Antibody-Producing Cell*" OR "Lymphocyte*" OR "Antigen-Presenting Cell*" OR "Dendritic Cell*" OR "Bone Marrow*" OR "Enterochromaffin*" OR "Immunological Synapse*" OR "Leukocyte*" OR "Granulocyte*" OR "Basophil*" OR "Eosinophil*" OR "Leukocyte*" OR "Mononuclear*" OR "Lymph*" OR "Mast Cell*" OR "Phagocyte*" OR "Macrophage*" OR "Monocyte*" OR "Neutrophil*" OR "B cell*" OR “lymphangiogene*” OR** “B-cell*” OR “T cell*” OR “T-cell*” OR “Cytokine*” OR “innate resp*” OR “innate react*” OR “autoimmun*” OR “Plasma cell*” OR “antibod*” OR “Major histocomp*” OR “Hapteni*” **) )** | 2,445 in 09/02/2024 |
| Embase | **('infant*':ti,ab,kw OR 'prenatal*':ti,ab,kw OR 'postnatal*':ti,ab,kw OR 'antenatal*':ti,ab,kw OR 'embryo*':ti,ab,kw OR 'fetal*':ti,ab,kw OR 'foetal*':ti,ab,kw OR 'foetus*':ti,ab,kw OR 'fetus*':ti,ab,kw OR 'gestation*':ti,ab,kw) AND ('agrochemical*':ti,ab,kw OR 'agrichemical*':ti,ab,kw OR 'plant protection product*':ti,ab,kw OR 'pesticid*':ti,ab,kw OR 'biocid*':ti,ab,kw OR 'herbicid*':ti,ab,kw OR 'weedkiller*':ti,ab,kw OR 'weed killer*':ti,ab,kw OR 'defolian*':ti,ab,kw OR 'insecticid*':ti,ab,kw OR 'nematicid*':ti,ab,kw OR 'molluscicid*':ti,ab,kw OR 'piscicid*':ti,ab,kw OR 'avicid*':ti,ab,kw OR 'rodenticid*':ti,ab,kw OR 'repellent*':ti,ab,kw OR 'antiparasit*':ti,ab,kw OR 'lampricid*':ti,ab,kw OR 'acaricid*':ti,ab,kw OR 'miticid*':ti,ab,kw OR 'mite control*':ti,ab,kw OR 'algicid*':ti,ab,kw OR 'algaecid*':ti,ab,kw OR 'chemosterilant*':ti,ab,kw) AND ('immun*':ti,ab,kw OR 'antibody-producing cell*':ti,ab,kw OR 'lymphocyte*':ti,ab,kw OR 'antigen-presenting cell*':ti,ab,kw OR 'dendritic cell*':ti,ab,kw OR 'bone marrow*':ti,ab,kw OR 'enterochromaffin*':ti,ab,kw OR 'immunological synapse*':ti,ab,kw OR 'granulocyte*':ti,ab,kw OR 'basophil*':ti,ab,kw OR 'eosinophil*':ti,ab,kw OR 'leukocyte*':ti,ab,kw OR 'mononuclear*':ti,ab,kw OR 'lymph*':ti,ab,kw OR 'mast cell*':ti,ab,kw OR 'phagocyte*':ti,ab,kw OR 'macrophage*':ti,ab,kw OR 'monocyte*':ti,ab,kw OR 'neutrophil*':ti,ab,kw OR 'b cell*':ti,ab,kw OR 'lymphangiogene*':ti,ab,kw OR '**B-cell***':ti,ab,kw** OR **'**T cell***':ti,ab,kw** OR **'**T-cell***':ti,ab,kw** OR **'**Cytokine***':ti,ab,kw** OR **'**innate resp***':ti,ab,kw** OR **'**innate react***':ti,ab,kw** OR **'**autoimmun***':ti,ab,kw** OR **'**Plasma cell***':ti,ab,kw** OR **'**antibod***':ti,ab,kw** OR **'**Major histocomp***':ti,ab,kw** OR **'**Hapteni***':ti,ab,kw)** | **997 in 09/02/2024** |
